# Use of Machine Learning Techniques for Predicting Heart Disease Risk from Phone Enquiries Data

**DOI:** 10.1101/2022.07.29.22278208

**Authors:** Fernando Martín-Rodríguez, Javier Pájaro-Lorenzo, Fernando Isasi-de-Vicente, Mónica Fernández-Barciela

## Abstract

This paper is about the application of known machine learning (ML) techniques for the prediction of heart disease risk. A public database is used to train and test the ML models. Results are evaluated using standard measures like precision, recall and F-score. ML models selected are well known techniques and they are based on different approaches. Chosen methods are: MLP (Multi-Layer Perceptron), SVM (Support Vector Machine) and Bagged Tree (Bootstrap Aggregated Trees). After evaluating techniques alone on their own, a new “triple voting method” (TVM) is tested applying the three individual methods and “adding” their results to improve accuracy.

## 1. Introduction

Artificial Intelligence (AI) is a discipline that, nowadays, is increasingly more used for many applications. Machine Learning (ML) is a core part of AI that is based on training mathematical models from revised examples from the real world. Models are, really, mathematical functions depending on numerical parameters. Training is an iterative method for adjusting parameters so that functions get the desired results on the given examples.

In our case, we will apply well known ML models to the problem of predicting the risk of heart disease from a reduced set of patient data.

To accomplish this, we will use the database known as “Heart-2020”: this database comes from the “2020 annual survey from the U.S. government’s CDC agency (Centers for Disease Control and Prevention)”. This is a public enquiry about the general state of citizens’ health. Originally, the survey was conducted by the Behavioral Risk Factor Surveillance System [1], considered the largest system of surveys on health of the planet, containing information from 279 variables in more than 400,000 patients. From the platform Kaggle [2,3], we can obtain freely an anonymized and reduced dataset related to heart disease probability. This is an annotated table (in the sense that we know if each individual has really developed a heart disease) with 17 fields (health data).

All data have been obtained enquiring the individuals, Id EST: we can obtain data for a patient through a simple interview, with no analysis or laboratory tests. So this kind of system can be useful to predict the number of heart disease problems in a given population and also to fire concrete alarms on given individuals, recommending them to follow further examinations and/or to improve some life habits.

In the literature, there are many other publications dealing with heart risk prediction [4,5,6], but they normally use more “difficult to collect” data. In this work we are restricting ourselves to the simple, easy to obtain, data from [1].

The fields (table columns) in this dataset are the following:

- BMI: Body mass index.
- Smoking: Yes/No.
- Alcohol Drinking: Yes/No.
- Stroke: Yes/No (has the individual suffered a stroke yet).
- Physical Health: how many days has the individual felt physically bad in the last 30.
- Mental Health: how many days has the individual felt mentally bad in the last 30.
- Difficulties Walking: Yes/No (has the individual difficulties for walking or for climbing stairs).
- Sex: biological sex: male/female.
- Age Category: 13 age intervals, starting at 18 years old.
- Race: 5 most common races and category “other”.
- Diabetic: Yes/No with special values “Borderline (near to diabetes)” and “During pregnancy”.
- Physical Activity: Yes/No (regular physical activity).
- General Health: poor/fair/good/very good/excellent.
- Sleep time: average sleep hours per day.
- Asthma: Yes/No.
- Kidney Disease: Yes/No (has the subject suffered a kidney disease).
- Skin Cancer: Yes/No (has the subject suffered from skin cancer).

In the remainder of the paper, we will describe the work with these data to train and test the models, as well as the obtained results.

## 2. Materials and Methods

### 2.1 Data Preparation

The main material for this work is the dataset that we have already described in the previous section. If we review the previously described fields, we can see that some of them are naturally numeric, for example: “BMI”. Other fields are binary (with a value of Yes or No), for example: “Smoking”. Finally, we have categorical fields; like, for example: “Race” or “Sex” (we consider Sex, categorical, not binary).

In order to apply our chosen methods, we need to convert all data into numeric. Binary data will be simply represented as “1” for “Yes” and “0” for “no”. For categorical fields, each column will be converted into several columns: one per category. Id EST: for example, “Sex” is divided into two binary fields: “Sex-Male” and “Sex-Female”, where each individual has a “1” value (and only one) in one of the new columns.

A special case is the field “Diabetic” that would be naturally binary except for the intermediate values: “Borderline” and “During Pregnancy”. In this case, we have decided to apply the following values: “0” (No), “0.50” (During Pregnancy), “0.75” (Borderline) and “1” (Yes).

A very important aspect of data preparation is the strong unbalancing of data in this dataset. Really, almost any dataset on “any” disease risk over a given population will be unbalanced because there will be much more “negative” cases than “positive” ones. Training in these conditions normally tends to useless systems because algorithm can lead to a system that “always says negative” because with that “functioning”, we would still obtain a high recognition rate (training methods are normally designed to achieve a high recognition rate or a minimum mean square error between achieved and desired output).

To correct unbalancing, we can use to approaches:

- Upsampling: “computing” new samples of the minority class (normally using interpolation).
- Undersampling: if we have enough number of samples, simply removing randomly samples of the majority class.

Any of the two techniques can lead to a bigger error rate in samples from the majority class when applied to real, non-balanced data. This is basically a bigger “false positive” probability, which in disease detection is not the worst problem.

After cleaning errors in the dataset, we have 319795 inputs (anonymized individuals), 27373 of them are positive (really suffered or are suffering heart disease). So we can downsample dataset to get a working set of 54746 individuals that is enough for our purposes.

### 2.2 Chosen methods

From authors’ experience in other ML projects, the following methods have been chosen:

#### MLP (Multi-Layer Perceptron)

This is a kind of artificial neural network [7] based normally on several stages of fully connected neuron layers. Sometimes weights are “pruned” so that some of the neurons can be “not fully connected”. Activation function is normally a “hyperbolic tangent” in the hidden layers and “sigmoid” or “softmax” in the output layer. MLP’s have proven to be very effective if there exists certain separability between classes. This means: “if feature vector clusters are physically separated”, although class frontiers can be geometrically complex. Note that in this problem, we already have a extracted feature vector. Then, there is no need to apply convolutional neural networks (CCN’s) that start by extracting high level features from low level data (pixels in an image, numerical samples of a ECG signal…).

#### SVM (Support Vector Machine)

This method[8] is based on translating the input feature vectors to “new vectors” in a greater dimensionality space. In that space, classes are linearly separable (this means using a hyperplane). SVM has been reported to have a similar accuracy to MLP’s except in the case of two classes, where SVM normally outperformsthe neural networks. This is exactly a “two classes problem”. In fact SVM classifiers for more than two classes are constructed using several two class SVM models.

#### BT (Bagged Tree or Bootstrap Aggregated Trees)

BT is a set of aggregated decision trees. When training, different subsets of data are created (not necessarily disjoint) to train different trees [9]. Those trees are run in parallel and prediction is averaged. BT is robust to noise or small variations of the input variables.

Finally, having tested three different methods with different approached and perhaps different strengths and weaknesses, we decided to test a joint method. Result from MLP is a single number between 0 and 1, with possible intermediate values (with decimal part). A result closer to 1 meansthat more probability of future disease is being predicted by the neural network for this given person.

Nevertheless, SVM and BT always give a “pure binary” prediction: “0” or “1”, there are no intermediate values. This can be seen as a drawback for these methods because we cannot quantify “certainty” of the system.

To join the three classifiers for a given individual, we simply add the three outputs (three numerical values) as three votes from three jury members. Knowing that one of the juries may produce a “non-categorical” result, we will consider a positive detection if the sum is over the “intermediate value”: 1.50. We call this method the “triple voting method” (TVM).

### 2.3 Feature Analysis

Feature analysis (or feature engineering) consists of assessing the importance of each feature vector component (field or table column), this can be used to simplify the feature vector and it can also improve performance if we get rid of non-relevant information.

After data preparation, we have 23 columns, that is not a very huge vector but testing some feature analysis methods can be worth if we get a better performance.

Within the available methods for feature selection, the ones based on feature filtering are the more interesting for us; because we can compute the importance of each feature prior to model building. Two of the most used methods are ANOVA and Pearson Correlation Coefficient.

#### ANOVA (Analysis of Variance)

basically the idea is that a feature is good when we split samples according to desired outputs and we get sets with different mean and small variance. A measure of importance is computed for each feature with mean differences in the numerator and a variance-like formula in the denominator [10].

#### Pearson Correlation Coefficient

it is a number between −1 and 1 computed from the cross correlation and variances of two random variables [10]. The greater the absolute value, the bigger relation between the two variables. Computing the correlation coefficient between each feature and the desired output can also be used as a measure of feature signification.

## 3. Results and Discussion

To assess accuracy of all methods we will put a part a randomly chosen (but balanced) subset of the (previously balanced) 54746 working samples. This subset will not be used for training and it will be saved for testing. It is a must to perform tests with samples that were not used for training.

We reserved 15% of samples for the test set. This means 8212 test samples, the rest: 54746-8212=46534 samples will be the training set.

Once trained, for the test set we compute the following numbers:

- TP: True positive (real positive samples classified correctly).
- TN: True negative (real negative samples classified correctly).
- FP: False positive (real negative samples classified incorrectly).
- FN: False negative (real positive samples classified incorrectly).

From this numbers, we can compute these measurements:

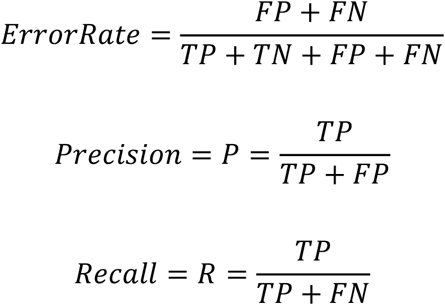

See that precision and recall give more information than simply the error rate. Remember that “saying always negative”, we perhaps will have a low error rate but zero precision and recall. What’s more, a high recall is perhaps the most important result when trying to detect risk of illness.

F-score is a combined number obtained from precision and recall. It is also called the “harmonic average”. It can be computed as “the inverse of the average inverse value”:

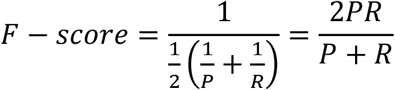

Obviously the greater F-score we achieve the better system we have. F will be a number between precision and recall but it is more demanding than arithmetic mean. F is nearer to the lowest value.

Other measure that we have computed is mean square error (MSE). This means considering the output of a method, over the entire test set, as a numerical vector (one vector component per individual output). This vector is compared with the “desired output vector” (1 for positive samples and 0 for negative ones). MSE is computed subtracting both vectors, adding the squares of components and dividing by length.

We will describe now the implementation details for each scheme. All the trainings and tests have been carried out in the MATLAB environment [11].

### 3.1 MLP

For the multilayer perceptron, we have to choose first network structure. We can speak about “meta-parameters” like number of layers, and number of hidden neurons.

For this kind of problem: a high level, not very large, feature vector and only two classes; the most simple structure is a net with only one output and only one hidden layer. In ML models simpler is usually synonym of “more appropriate” (it is good idea to avoid very complex structures that can run into learning problems). In these conditions, the chosen structure is this one:

Unfortunately there is no established method for computing the size of the hidden layer. Normally it is determined empirically testing intermediate values between the number of inputs and the number of outputs (minimum two hidden neurons, only one would be nonsense). In our case, 10 yielded good results.

The training algorithm we used is the most popular: backpropagation with conjugate gradient. Besides the 15% samples reserved for final testing, algorithm saves other 15% for validation (for deciding when to stop the algorithm).

Results can be inferred from the following figure (confusion matrix):

Where numbers in bold are true counts of test samples in each category. Columns are organized according to the desired result (target) so that, for example: first column is made of true negative samples, the box in green counts the ones correctly classified and the box in pink the ones incorrectly classified.

Measurements for MLP would be as follows.

Error rate: 0.23

Precision: 0.75

Recall: 0.80

F-score: 0.78

MSE: 0.23

Having a high recall and F-score, we can consider it a promising result.

### 3.2 SVM

SVM was trained in MATLAB using standard tools. Kernel function is linear and the optimization method is ISDA (Iterative Single Data Algorithm).

Numerical results were as follows.

Error rate: 0.24

Precision: 0.74

Recall: 0.80

F-score: 0.77

MSE: 0.24

Being results very similar to those of the MLP.

### 3.3 Bagged Tree

Here the main meta-parameter is the number of trees to be aggregated. We started with 10 trees and then we increased this value, reaching even the number of 100 trees. We got very similar results in all the tests, so we returned to 10 trees.

We know present the numerical results in the same manner of the former options.

Error rate: 0.26

Precision: 0.73

Recall: 0.77

F-score: 0.75

MSE: 0.26

Which is slightly worse than the two former options.

### 3.4 Triple Voting

Having three different methods with so similar capabilities encourages the idea of getting a better system combining strengths from all approaches.

Details for this combined method (TVM: Triple Voting Method) have are already been explained in section 2. We have repeated the test with the same samples, obtaining the following.

Error rate: 0.23

Precision: 0.75

Recall: 0.81

F-score: 0.78

MSE: 0.23

It seems as good as the former methods but we do not find a significant improvement. Anyway, by caution, TVM should be chosen as the most robust method, because it is using the potential of all the individual ones. As execution time is not heavy for ML methods (training can be very time consuming but at run time are normally fast), real-time issues are not a barrier for TVM here.

What happens if we test with the whole (non-balanced database)? It is expectable that with majority of negative samples, we get an important number of false positive results that make precision to get worse. Numerical results are listed below.

Error rate: 0.26

Precision: 0.22

Recall: 0.81

F-score: 0.35

MSE: 0.26

We see that precision goes dramatically down pulling F-score accordingly. The rest of parameters are almost maintained. Having into account that recall has a very good value this is not a so bad result. In fact, this problem arises with any of the individual methods. Testing with the whole database, all parameters are approximately the same, except for precision that goes dramatically down (MLP: 0.22, SVM: 0.22, BT: 0.24) and F-score that diminishes accordingly (MLP: 0.34, SVM: 0.34, BT: 0.38).

### 3.5 Application of Feature Engineering

Computing the ANOVA quotient values and the correlation coefficients for all the given features (as described in section 2.3), we obtain the following values, see curves in figure 3: blue for ANOVA quotient (normalized) and red for correlation coefficient (absolute value).

**Fig. 1.**
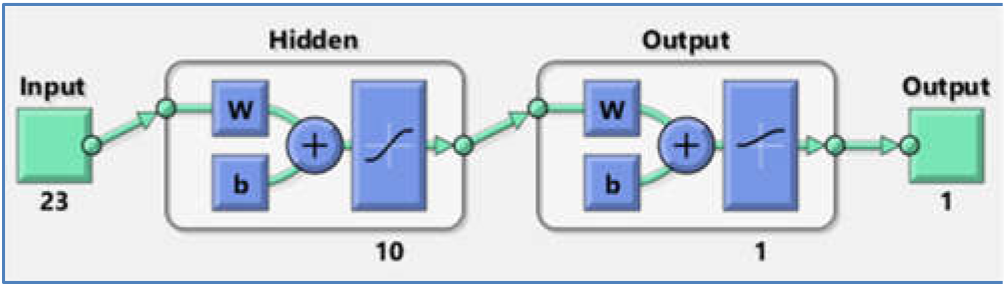
MLP structure.

**Fig. 2.**
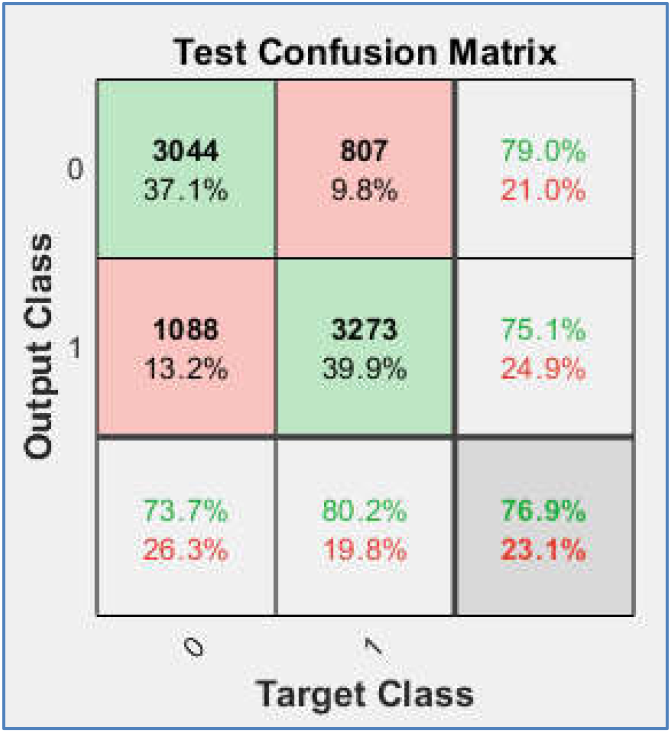
Confusion matrix for the TEST set.

**Fig. 3.**
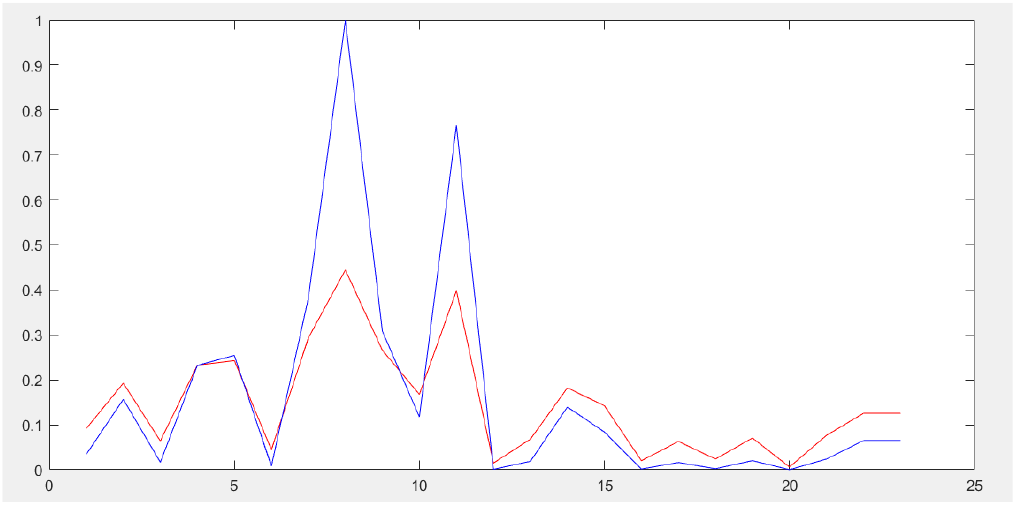
Confusion matrix for the TEST set.

Obviously value ranges are different but we can see that local maxima coincide, except for the last set of features. These last features are the columns that were expanded for the categorical features: race and biological sex. These sets of columns should be considered as a whole, where all are included or all are discarded.

Race seems not very significant according to values in both curves and sex is going to be taken into account. For these reasons we are going to discard expanded columns in this study.

Note also that Pearson coefficient could be less reliable in this problem because output is binary and this case is better handled by ANOVA technique. Nevertheless, figure 3 demonstrates that both measurements basically coincide.

Selecting the features with significant values in these curves (greater than 20% percent of maximum value in ANOVA coefficient), we test a new feature vector with only SIX features. The selected features are: “Stroke (Yes/NO)”, “Physical Health”, “Difficulties Walking”, “Age Category” and “Physical Activity”.

All methods have been retrained and tested with this reduced dataset. Results have been very similar to the previous ones but always slightly worse. As example, we summarize numbers for the joint (TVM) method:

Error rate: 0.24

Precision: 0.74

Recall: 0.80

F-score: 0.77

MSE: 0.24

As the total number of features (23) is not excessive, we conclude that feature selection is not necessary in this case, although it could be used if we want to create a system that gives a response with very few questions.

### 3.6 Splitting by Gender

Knowing that heart stroke symptoms are totally different for men and women, we thought that perhaps splitting the dataset by gender could improve results. That means training two different systems: one for men and other for women using two separate datasets where we will have removed the “biological sex” columns.

We proceeded, finding that we have 151990 men and 167805 women in the original (non-balanced database). We also have 16139 cases of positive heart disease for men and 11234 for women. “A priori” probability for heart disease, according to this data, would be less for women: 10.62% vs. 6.69%.

Using the same method for balancing the two datasets, we will have two independent matrices of 32278 samples (men) and 22468 samples (women). Running the same tests for these two new sets, we get the following results; we have summarized them in a table using the “recall” and “F-score” values for comparison:

**Table I.**
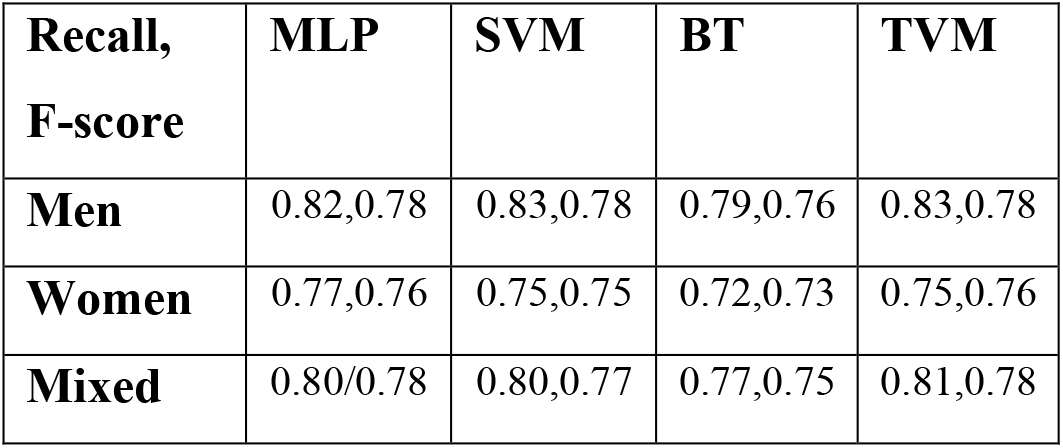
Comparison of results.

From the table, we can see that results improve a bit for men and get a bit worse for women. Perhaps this can be due to the less number of positive cases between women that results in a smaller training set.

Facing these data our recommendation would be to use a separate system, knowing that “further study” recommendation will be stronger in the case of men. Of course this opens a future line in order to get more balanced results.

### 3.6 Conclusions and Future Lines

We have designed a system for predicting heart disease risk using ML techniques and easily measurable parameters that can be obtained by patient itself or via telephone or internet enquiries. Results are promising reaching a F-score of 0.78 on balanced datasets. Feature vector reduction is possible but not necessary and, in this case, produces no performance improvement.

System can be used “as is” for individual advice to patients in order to make recommendations about further medical tests and/or changes in lifestyle. Note that enquiries can be performed by an online form so that individuals can “auto-test” themselves. Gender splitting can be adequate for this purpose, knowing that with present database, results are more accurate for men.

If we have massive amounts of data taken from the general population, this system can be used for generating estimations of future heart disease rate and to take the appropriate preventive actions.

Future lines, today, would be:

- Testing ML algorithms with other configurations (meta-parameters) trying to improve results.
- Testing new ML algorithms and integrating them in the combined (voting) scheme.
- Getting more gender balanced results in the case of gender splitting.
- Applying this methodology to other databases of interest for public health.

## Data Availability

All data produced in the present study are available upon reasonable request to the authors.

https://www.kaggle.com/datasets/kamilpytlak/personal-key-indicators-of-heart-disease/discussion/316999?datasetId=1936563

